# Genetic Susceptibility to Atrial Fibrillation Identified via Deep Learning of 12-lead Electrocardiograms

**DOI:** 10.1101/2022.01.17.22269357

**Authors:** Xin Wang, Shaan Khurshid, Seung Hoan Choi, Sam Friedman, Lu-Chen Weng, Christopher Reeder, James P. Pirruccello, Pulkit Singh, Emily S. Lau, Rachael Venn, Nate Diamant, Paolo Di Achille, Anthony Philippakis, Christopher D. Anderson, Jennifer E. Ho, Patrick T. Ellinor, Puneet Batra, Steven A. Lubitz

## Abstract

Artificial intelligence (AI) models applied to 12-lead electrocardiogram (ECG) waveforms can predict atrial fibrillation (AF), a heritable and morbid arrhythmia. We hypothesized that there may be a genetic basis for ECG-AI based risk estimates. We applied an ECG-AI model for predicting incident AF to ECGs from 39,986 UK Biobank participants without AF. We then performed a genome-wide association study (GWAS) of the predicted AF risk. We identified three signals (*P*<5×10^−8^) at established AF susceptibility loci marked by the sarcomeric gene *TTN*, and sodium channel genes *SCN5A* and *SCN10A*. We also identified two novel loci near the genes *VGLL2* and *EXT1*. In contrast, a GWAS of risk estimates from a clinical variable model indicated a different genetic profile. Predicted AF risk from an ECG-AI model is influenced by genetic variation implicating sarcomeric, ion channel, and height pathways. ECG-AI models may identify individuals at risk for disease via specific biological pathways.

## Introduction

Atrial fibrillation (AF) is a heritable arrhythmia associated with substantial morbidity, including stroke, heart failure, dementia, and mortality.^1–3^ Identifying individuals at high risk of developing AF may enable early detection via cardiac rhythm monitoring and treatment, or behavioral modification to prevent AF altogether. Artificial intelligence (AI) algorithms applied to 12-lead electrocardiogram (ECG) waveforms can predict AF.^4–6^ Algorithms that predict AF risk from ECGs have practical appeal given the ubiquity and inexpensive nature of ECGs, and lack of requirement for manual data input for risk estimation. Whether risk estimates derived from AI algorithms reflect specific underlying genetic pathways that increase susceptibility to AF is unclear.

Understanding the biological basis for risk estimates from machine learning models could aid model interpretability, rationalize model outputs, promote clinician confidence, and potentially enable identification of individuals with specific mechanistic pathways that lead to AF. We recently developed and validated an AI algorithm for predicting the 5-year risk of new-onset AF using 12-lead ECGs (“ECG-AI”).^7^ In the present study, we conducted genetic association testing with AF risk estimates generated from the ECG-AI model to assess the genetic underpinnings reflected by the output. As a comparator, we assessed the genetic basis of a widely validated clinical risk factor model for predicting AF, the CHARGE-AF score (Cohorts for Aging and Research in Genomic Epidemiology–AF).^8^

## Methods

### Study sample

Individual-level data used in the present study were from the UK Biobank, a prospective cohort study comprising 502,629 individuals aged 40 to 69 at enrollment from the United Kingdom during 2006-2010.^9,10^ Briefly, phenotypic data comprising survey, longitudinal electronic health record, national registry, imaging, and other bioassay data were collected. In total 488,377 participants underwent genome-wide genotyping as summarized below. Of these, 39,986 individuals who underwent 12-lead ECG collection and had complete data to estimate the CHARGE-AF score, were without a history of AF at the time of ECG acquisition, and in whom data passed quality control (QC) procedures, were included in the genome-wide association studies (GWAS) of predicted AF risk using either ECG-AI or CHARGE-AF. Of the remaining 448,391 individuals without ECGs, 424,411 that passed genome-wide genotyping QC procedures were included in polygenic risk score analyses. The study overview and analytic workflow can be found in **Figure 1**. All UK Biobank participants provided electronic signed consent at recruitment, and the study protocol was approved by the UK Biobank Research Ethics Committee (reference number 11/NW/0382). Use of data (under UK Biobank application 7089) for the current study was approved by the Mass General Brigham (MGB) Institutional Review Board.

**Figure 1.**
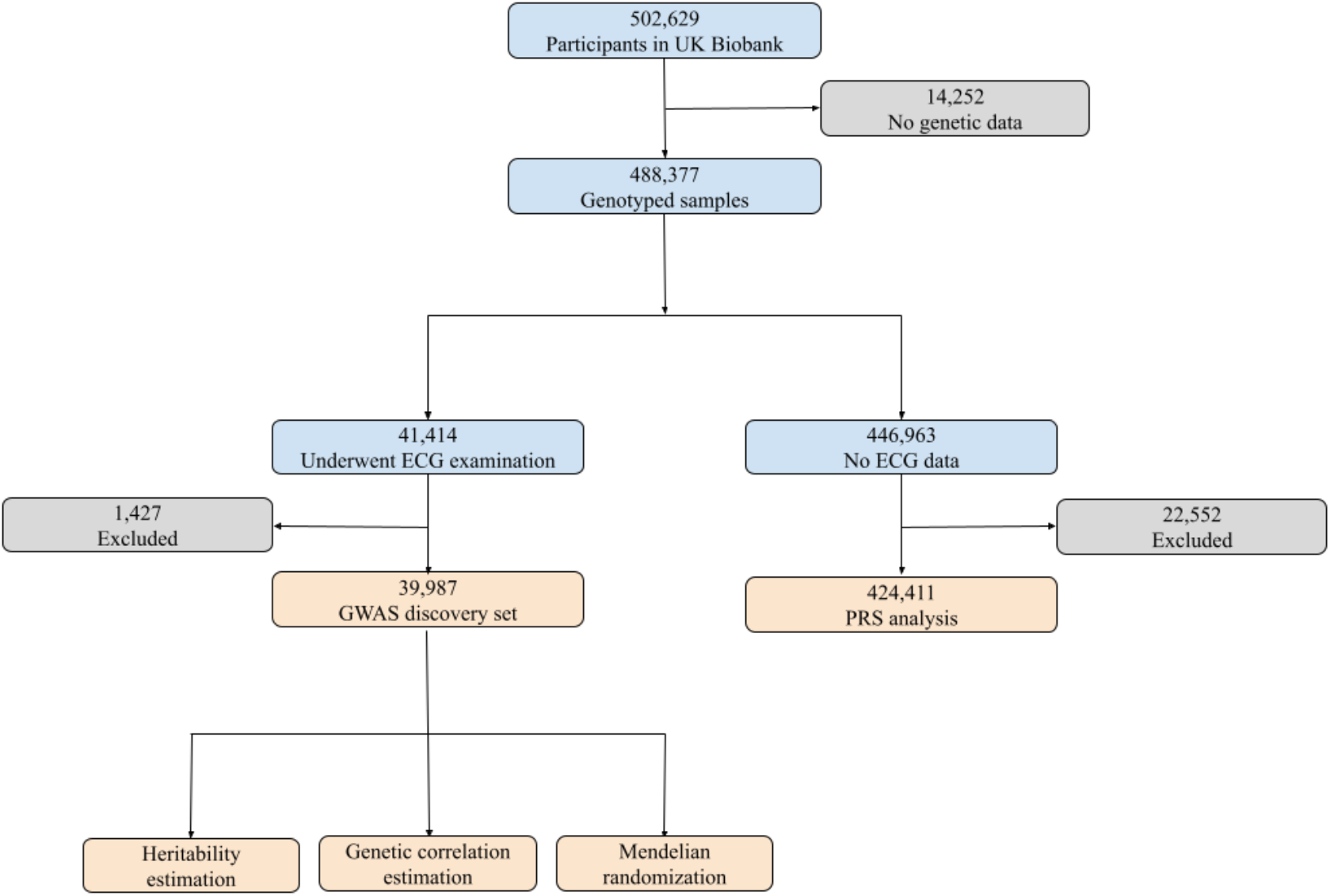
Study overview. We applied our validated ECG-AI model to samples with ECG data in the UK Biobank. After excluding participants who withdrew consent, had missing CHARGE-AF components, were diagnosed with atrial fibrillation (AF) before ECG examination, did not have follow-up information or failed sample QC procedures, 39,987 remained in the discovery set, in which we performed GWAS and post-GWAS analyses. Among the 446,963 remaining genotyping samples, 424,411 did not withdraw consent, were not < 3rd-degree relatives with individuals in the GWAS set, were not diagnosed with AF before enrollment, had follow-up information, and did not fail sample QC procedures. We calculated polygenic risk scores (PRS) using the GWAS results and associated the PRS with incident AF in this subset.

### Predicted risk of AF

Our study outcome was the predicted 5-year risk of AF. The 5-year risk of AF was derived from both an ECG-AI model, and separately from the CHARGE-AF clinical risk factor model. The development and validation of the ECG-AI model has previously been reported.^7^ Briefly, we trained a convolutional neural network (CNN) to predict 5-year risk of AF using 12-lead ECGs in a longitudinal patient cohort derived from the MGB network.^11^ ECG-AI uses an encoding and loss function that takes into account both time to event (i.e., AF) and missingness introduced by censoring (death or loss of follow-up) to estimate a 5-year survival probability of AF. The predicted AF risk was then calculated as one minus the survival directly outputted from the ECG-AI model. We assessed discrimination of ECG-AI predicted risk for 5-year AF by calculating Harrell’s c-index^12^ and found a moderate discrimination of 0.70 (95% confidence interval: 0.67-0.72) in the 39,986 GWAS discovery set.

We calculated the CHARGE-AF predicted 5-year risk of AF using the published equation 1-0.9718412736^exp(*X*-12.5815600)^, where *X* is the CHARGE-AF score for each individual.^8^ Clinical risk factors in the score include baseline age, race, height, weight, systolic and diastolic blood pressure, smoking status, antihypertensive medication use, diabetes, heart failure, and myocardial infarction. Clinical variables were based on self-report (race, smoking status, antihypertensive medication use), measurements at the ECG visit in the UK Biobank (age at ECG, height, weight, systolic and diastolic blood pressure), or inpatient international classification of diseases (ICD) 9 or 10 codes (diabetes, heart failure, myocardial infarction) as has been described previously.^7^ CHARGE-AF has been validated widely and shown to have good discrimination with C-statistics ranging from 0.66 to 0.81.^8,13,14^ The ECG-AI and CHARGE-AF score linear predictors have previously been reported to have modest correlation (Pearson r 0.66) in an independent dataset, and the CHARGE-AF score exhibits similar predictive performance as ECG-AI.^7^

For survival analysis in the present study, incident AF at 5 years was ascertained using follow-up information and a previously published definition comprising inpatient ICD-9 and -10 codes, and procedural codes.^15^ Follow-up information in the UK Biobank depends on the availability of linked electronic health records and hence varies by enrollment sites. Censoring occurred at death, withdrawal of consent, or last availability of hospital linked data.^7^

### Genetic data

Genotyping was carried out using two different arrays (UK BiLEVE Axiom Array and UK Biobank Axiom Array) with 95% shared marker content. After variants and sample QC processes, these markers were phased and imputed to the Haplotype Reference Consortium (HRC) and the merged UK10K and 1000 Genomes phase 3 reference panels, resulting in ∼96 million variants in the final dataset.^10^ We used the QC metrics provided by the UK Biobank to perform sample QC. Participants who had mismatch between self-reported sex and genetically inferred sex, high genotype missingness rate, high heterozygosity rate, or sex chromosome aneuploidy were removed from the analysis. We used genotyped markers to fit null models for the GWAS. Variants were filtered out if MAF was < 0.1%, minor allele counts (MAC) was < 100, missing call rate was > 0.1, or Hardy-Weinberg equilibrium exact test *P* < 1×10^−5^. In addition, samples with a missingness call rate > 0.1 were excluded from the analysis. We then performed the association tests using imputed genetic data, and variants included were those with a minor allele frequency (MAF) > 0.1% and an information score (imputation quality) > 0.3.

### Common variant analyses

We conducted a GWAS of ECG-AI predicted 5-year AF risk, and a separate GWAS of CHARGE-AF predicted 5-year AF risk as a comparator. Rank-based inverse normal transformation (R-INT) was applied to each phenotype before association testing. We used the software Regenie,^16^ which implements an efficient whole-genome regression approach, for this analysis assuming an additive genetic model for each single nucleotide variant (SNV).

Covariates adjusted in statistical models included age at the time of ECG, sex, genotyping array, and the first 20 principal components of ancestry. We also conducted conditional analyses^17^ using the GCTA software to select secondary independent significant SNVs at each locus. In addition, as a comparator, we performed a GWAS of observed 5-year incident AF in the same dataset using Cox Mixed-Effects models^18^ implemented in the *coxmeg* R package with the same covariates. Due to computational burden, we only tested variants with MAF > 10% and information score (imputation quality) > 0.9. Genome-wide significant variants were considered those with *P* < 5×10^−8^. The mapped gene at each risk locus was either the nearest gene or the trait-associated gene within 500 kb of the lead SNV reported in the GWAS catalog.

### Heritability and genetic correlation

We calculated the additive heritability (*h*^*2*^) for predicted risk of AF using a variance-components analysis with individual-level genetic and phenotypic data in our GWAS set, adjusting for age at the time of ECG, sex, genotyping array, and the first 20 principal components of ancestry using BOLT-REML.^19^ We also used the same method to estimate the genetic correlation between the GWAS of predicted AF risk and 5-year observed incident AF. BOLT-REML implements a Monte Carlo average information restricted maximum-likelihood algorithm to estimate the genetic variance-covariance matrix. We then utilized GWAS summary statistics with LD score regression (LDSC)^20^ to corroborate the heritability and genetic correlation estimates. GWAS results of predicted AF risk from the above common variant analyses were compared to a prior large-scale GWAS of AF.^21^ We used pre-computed LD-scores provided by the LDSC software package. Here we report the raw genetic correlation estimates without any conversion since these parameters accurately reflect the genetic correlation of underlying disease liabilities.^22^ Heritability estimates were also not converted from observed scale to liability scale since predicted risks were normally distributed continuous traits after rank-based inverse normal transformation.

### Polygenic risk scores

We derived polygenic risk scores (PRS) for ECG-AI and CHARGE-AF predicted risk of AF using the GWAS results and then applied these to the remaining participants without ECG data (**Figure 1**). In addition to the sample QC procedures described above, we further excluded participants with > 3rd degree relatedness in the GWAS set. The SNV effect sizes used in score calculations were obtained by using a fully Bayesian approach, PRS-CS,^23^ which applies shrinkage factors to the original GWAS effect sizes to avoid overfitting. The polygenic score was calculated as the PRS-CS adjusted SNV effect size-weighted sum of the individual’s genetic dosage at effect alleles. Score calculations were carried out using PLINK 2.0 [https://www.cog-genomics.org/plink/2.0/].

### Associations between polygenic risk scores and incident AF

We tested the association between each polygenic risk score and 5-year incident AF using a Cox proportional hazards model adjusting for age at enrollment, sex, genotyping array, and the first 20 principal components of ancestry. We also fit another model containing both PRSs with the same covariates to estimate the effect of each PRS on 5-year incident AF risk after conditioning on the other PRS. We then further extended the model to include an interaction term between the two PRSs to assess for effect modification. We plotted the cumulative incidence of AF stratified by distribution of PRS risk (highest 10%, middle 80%, and lowest 10%). We performed the above analyses in the entire testing set as well as a subset of White British participants to assess the properties of PRSs across ancestries. All analyses were conducted using *survival* and *survminer* packages in R.

### Mendelian Randomization

We employed a two-sample Mendelian randomization (2SMR) approach^24^ implemented in the *TwoSampleMR*^25^ R package to estimate potential causal effects of exposures on outcomes using GWAS summary statistics. Genetic instruments were chosen from GWAS results of exposure traits if they reached *P* < 5×10^−8^. A clumping step was then performed to select independent variants within 10,000 kb distance with a maximum LD r^2^ of 0.001. These SNPs were then extracted from our ECG-AI risk summary statistics if they were present in both GWAS. The resulting genetic instruments were harmonized before analysis to ensure the effect alleles were consistent between the two studies. Summary statistics for P wave duration^26^ (unit: ms) and height^27^ (unit: SD) were chosen from studies that did not include UK Biobank samples. For each exposure, we performed MR analysis using five methods, including inverse variance weighted approach, weighted median approach, MR-Egger,^28^ MR-PRESSO,^29^ and MR-PRESSO corrected for horizontal pleiotropic outliers.

## Results

### Sample characteristics

The mean age of the 39,986 participants included in the GWAS was 64.0 +/-7.7 years at the time of ECG acquisition and 52% were female. The median follow-up time (starting from ECG visit) for this subset was 2.8 years (quartile 1: 1.9, quartile 3: 4.3). 510 individuals developed incident AF within 5 years of follow-up, corresponding to a cumulative incidence of AF of 2.12%. The mean age of the 424,411 participants included in the polygenic risk score application analysis who did not have ECGs was 57.1 +/-8.1 years at the time of study enrollment and 55% were female. The median follow-up time (starting from study enrollment) for this subset was 11.1 years (quartile 1: 10.4, quartile 3: 11.8). 7,077 individuals developed incident AF within 5 years of follow up, corresponding to a cumulative AF incidence of 1.70%. Participant characteristics are presented in **Table 1**.

**Table 1.** Characteristics of the UK Biobank cohort participants. Clinical variables were ascertained at baselines. For the genome-wide association studies (GWAS) discovery set, baseline refers to the time of electrocardiogram (ECG) visit. For the polygenic risk scores (PRS) testing set, baseline refers to the time of study enrollment. Descriptive statistics of enrollment age, female, race, and history of diabetes, heart failure and myocardial infarction for the PRS testing set were calculated using the complete sample (N=424,411). Height, weight, systolic blood pressure and diastolic blood pressure were summarized in a subset with the data available (N=423,044). The sample size for participants who self-reported their smoking status and medication use were 423,919 and 477,644, respectively.

| **Variable** | **GWAS set**<br>**(N=39,986)** | **PRS testing set**<br>**(N=424,411)** |
| --- | --- | --- |
| **Enrollment age** | 55.6 (7.6) | 57.1 (8.1) |
| **ECG age** | 64.0 (7.7) | Not applicable |
| **Female** | 20,809 (52.0%) | 232,654 (54.8%) |
| **Race (White British)** | 38,617 (96.6%) | 370,121 (87.2%) |
| **Height (cm)** | 169.1 (9.2) | 168.3 (9.3) |
| **Weight (kg)** | 76.0 (15.2) | 78.0 (16.0) |
| **Systolic blood pressure** | 138.3 (18.6) | 138.1 (18.7) |
| **Diastolic blood pressure** | 79.1 (10.1) | 82.4 (10.2) |
| **Smoking (current)** | 1,446 (3.6%) | 46,433 (11.0%) |
| **Antihypertensive medication use (Yes)** | 4,275 (10.7%) | 88,887 (18.6%) |
| **Diabetes (Yes)** | 1,546 (3.9%) | 11,089 (2.6%) |
| **Heart failure (Yes)** | 186 (0.5%) | 1,701 (0.4%) |
| **Myocardial infarction (Yes)** | 916 (2.3%) | 9,657 (2.3%) |

### Genome-wide association analyses

The GWAS of ECG-AI predicted AF risk did not demonstrate any inflation (λgc=1.04). Four genome-wide significant (*P* < 5×10^−8^) loci were identified (**Figure 2** and **Supplementary Table** Two of the top SNVs were in close proximity to genes previously reported to be associated with AF, including *TTN* and *SCN10A*.^21,30^ The nearest genes at the other two loci were *VGLL2* and *EXT1*. A conditional analysis detected an additional independent association signal at the *SCN5A* locus (**Supplementary Table 1**), which has also been reported to be associated with AF in previous studies.^21,31^

**Figure 2.**
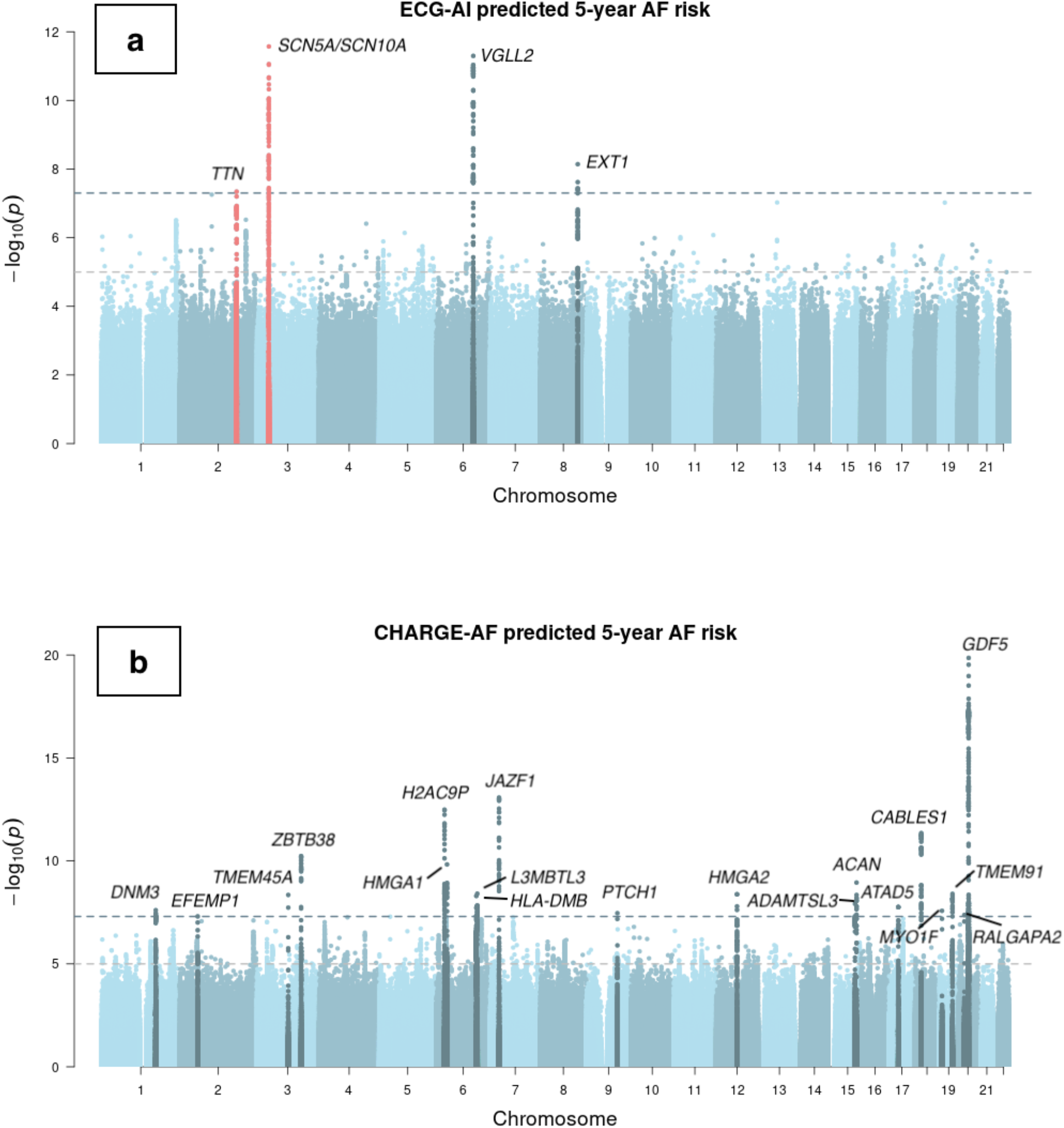
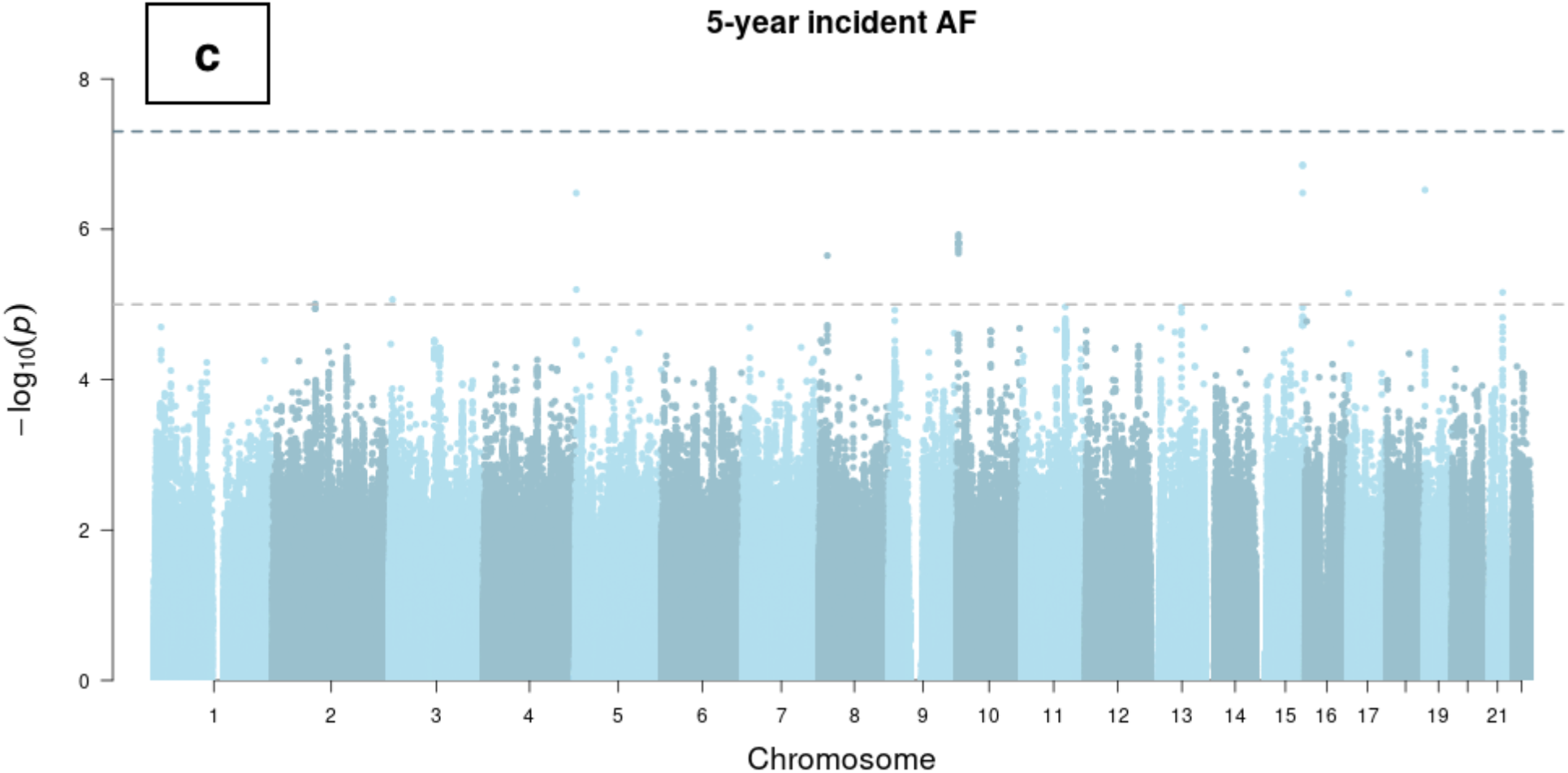
Manhattan plots of genome-wide association studies of ECG-AI and CHARGE-AF predicted risk of AF, and observed 5-year incident AF in the UK Biobank. Chromosomal variant positions are plotted on the x-axis. The -log10(*P* values) are plotted on the y-axis. The genome-wide significance threshold (5×10^−8^) is indicated by the horizontal dotted line. Variants are colored red near loci that have been reported in a prior atrial fibrillation (AF) GWAS,^21^ and are colored dark blue near loci that have not been reported previously in association with AF. Panels display associations with (a) ECG-AI predicted 5-year risk of AF, (b) CHARGE-AF predicted 5-year risk of AF, and (c) observed incident AF at 5-years in the UK Biobank. 5-year AF risk estimates were rank-based inverse normal transformed prior to analysis (see text).

In the GWAS of CHARGE-AF predicted risk, minimal genomic inflation was observed (λgc=1.10) which was likely due to polygenicity rather than population stratification, as implicated by the LD score regression intercept (1.0107). Nineteen loci were identified in the GWAS of CHARGE-AF predicted risk (**Figure 2** and **Supplementary Table 2**), none of which have previously been reported in association with AF. Traits associated with lead SNVs at these loci mainly consist of body size measurements and phenotypes related to the clinical factors included in the CHARGE-AF score calculation (**Supplementary Table 2** and **Supplementary Table 3**). No secondary association signals were detected in a conditional analysis.

The LocusZoom plots for risk loci identified in the two GWAS were presented in **Supplementary Figure 1** and **Supplementary Figure 2**, respectively. We also compared the summary statistics of the independent significant lead SNVs in the ECG-AI risk GWAS to that in the CHARGE-AF risk GWAS (**Supplementary Table 4**). No variants exceed the genome-wide significance threshold in the GWAS of 5-year incident AF with the same covariates (**Figure 2**).

### Heritabilities and genetic correlations

Using individual level genomic and phenotypic data, the estimated heritability (*h*^*2*^) was 13.0% (s.e. 1.4%) for ECG-AI risk and 36.5% (s.e. 1.4%) for CHARGE-AF risk. Genetic correlations with AF were estimated to be 35.3% (s.e. 13.7%) for ECG-AI risk and 18.9% (s.e. 8.6%) for CHARGE-AF risk. We further estimated the genetic correlation between the predicted AF risks from ECG-AI and CHARGE-AF, and found a significant correlation of 39.3% (s.e. 4.5%). As a comparator, we also calculated the heritabilities and genetic correlations using GWAS summary statistics with LD score regression,^20^ and the estimates were similar in magnitude to those calculated from individual-level data. Detailed results are provided in **Figure 3** and **Supplementary Table 5**.

**Figure 3.**
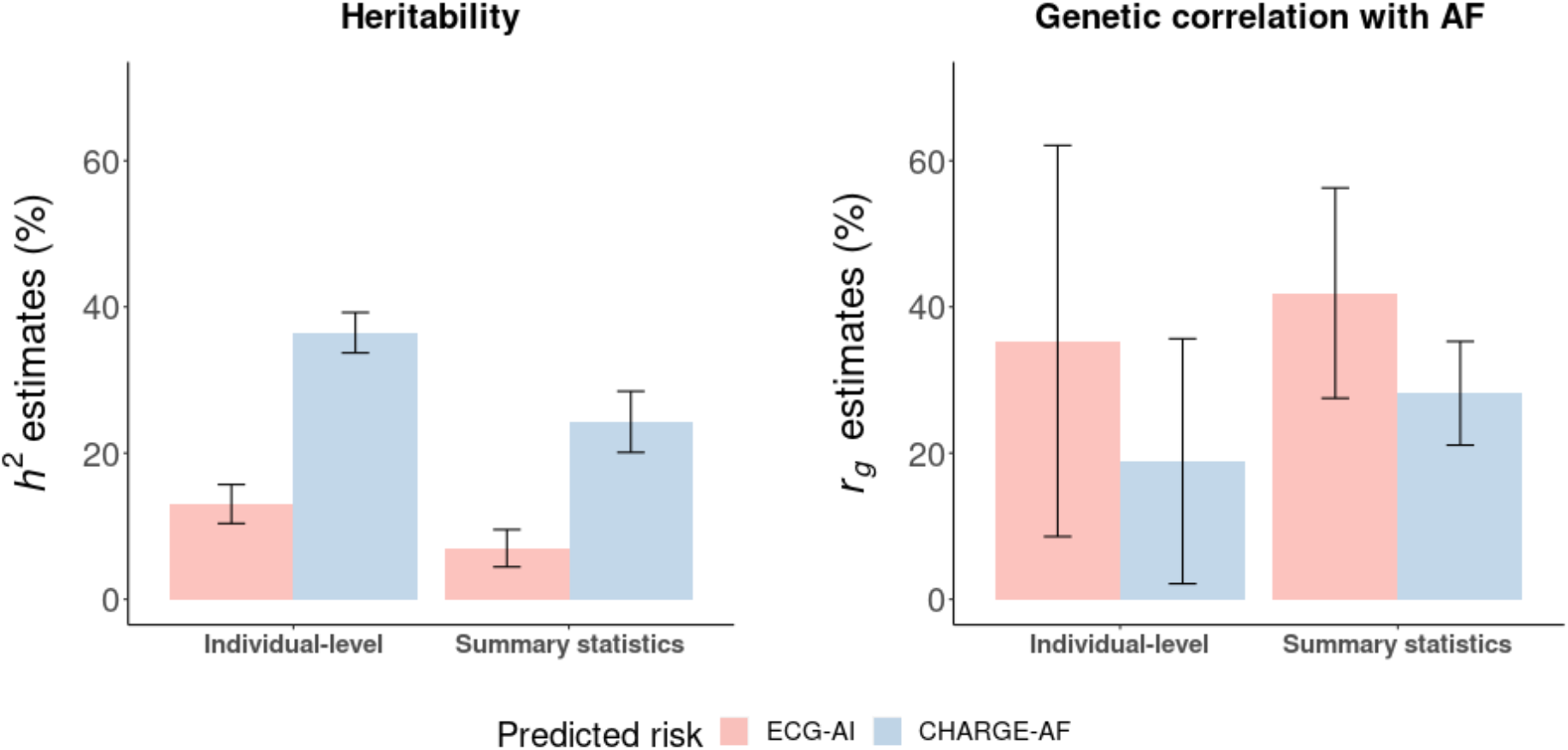
Heritability and genetic correlation estimates for model predicted 5-year atrial fibrillation (AF) risk. Heritability (*h*^*2*^) derived from the ECG-AI and CHARGE-AF GWAS are displayed on the left panel. Genetic correlation (*r*_*g*_) comparing both the ECG-AI and CHARGE-AF GWAS with a prior independent large-scale GWAS of AF,^21^ is displayed on the right panel. ‘Individual-level’ refers to estimates generated from individual-level genetic and phenotypic data. ‘Summary statistics’ refers to estimates generated from GWAS results. Summary statistics for ECG-AI risk and CHARGE-AF risk were extracted from the GWAS in the present study.

### Polygenic risk scores and incident AF

We calculated two polygenic risk scores (PRS) for the 424,411 eligible participants using predicted AF risk GWAS results. Each 1-SD increase in the PRS of R-INT ECG-AI risk (PRS_ECG-AI_) and the PRS of R-INT CHARGE-AF risk (PRS_CHARGE-AF_) were significantly associated with 5-year incident AF (PRS_ECG-AI_ hazard ratio [HR] 1.07, 95% CI 1.04 - 1.09, *P* = 3.0×10^−8^; and PRS_CHARGE-AF_ HR 1.12, 95% CI 1.09 - 1.14, *P* = 3.4×10^−19^). When included in the same model, both remained significantly associated with 5-year incident AF (PRS_ECG-AI_ HR 1.06, 95% CI 1.04 - 1.09, *P* = 1.1×10^−6^; and PRS_CHARGE-AF_ HR 1.11, 95% CI 1.08 - 1.14, *P* = ×10^−17^). We did not observe a significant interaction between the two PRSs (*P* = 0.97). We also plotted the cumulative risk of AF stratified by high (10%), middle (80%), and low (10%) groupings of the PRS distributions and observed separation between groups (**Supplementary Figure 3**). Due to a difference in the ancestral composition of the GWAS discovery set (White British: 96.6%) and PRS testing set (White British: 87.2%), we repeated the above analysis in the subset of the PRS testing set comprising White British participants only. We observed similar results (**Supplementary Table 6** and **Supplementary Figure 4**).

### Contributing components of ECG-AI predicted AF risk

Informed by the genetic signals from our common variant analysis, we tested the causal effects of P wave duration and body height on ECG-AI predicted risk using a two sample Mendelian Randomization (MR) approach. We extracted GWAS summary statistics from studies that did not include UK Biobank samples for P wave duration and body height (see **Methods**). The P wave was selected because (1) it has been linked to *SCN5A/SCN10A* in previous genetic studies,^26^ and (2) it had the greatest impact on ECG-AI predicted risk indicated by saliency maps and a median waveform analysis in our previous study.^7^ Body height was selected because (1) it has been linked to *VGLL2* and *EXT1* loci,^32,33^ and (2) is an established risk factor for AF.^34,35^

A significant and plausible causal effect of P wave duration on ECG-AI risk was supported by four out of the five methods we used, with MR effect sizes ranging from 0.017 to 0.023. The causal effect of height on ECG-AI risk was supported by all five methods, with MR effect sizes ranging from 0.085 to 0.123. Results are presented in **Figure 4**.

**Figure 4.**
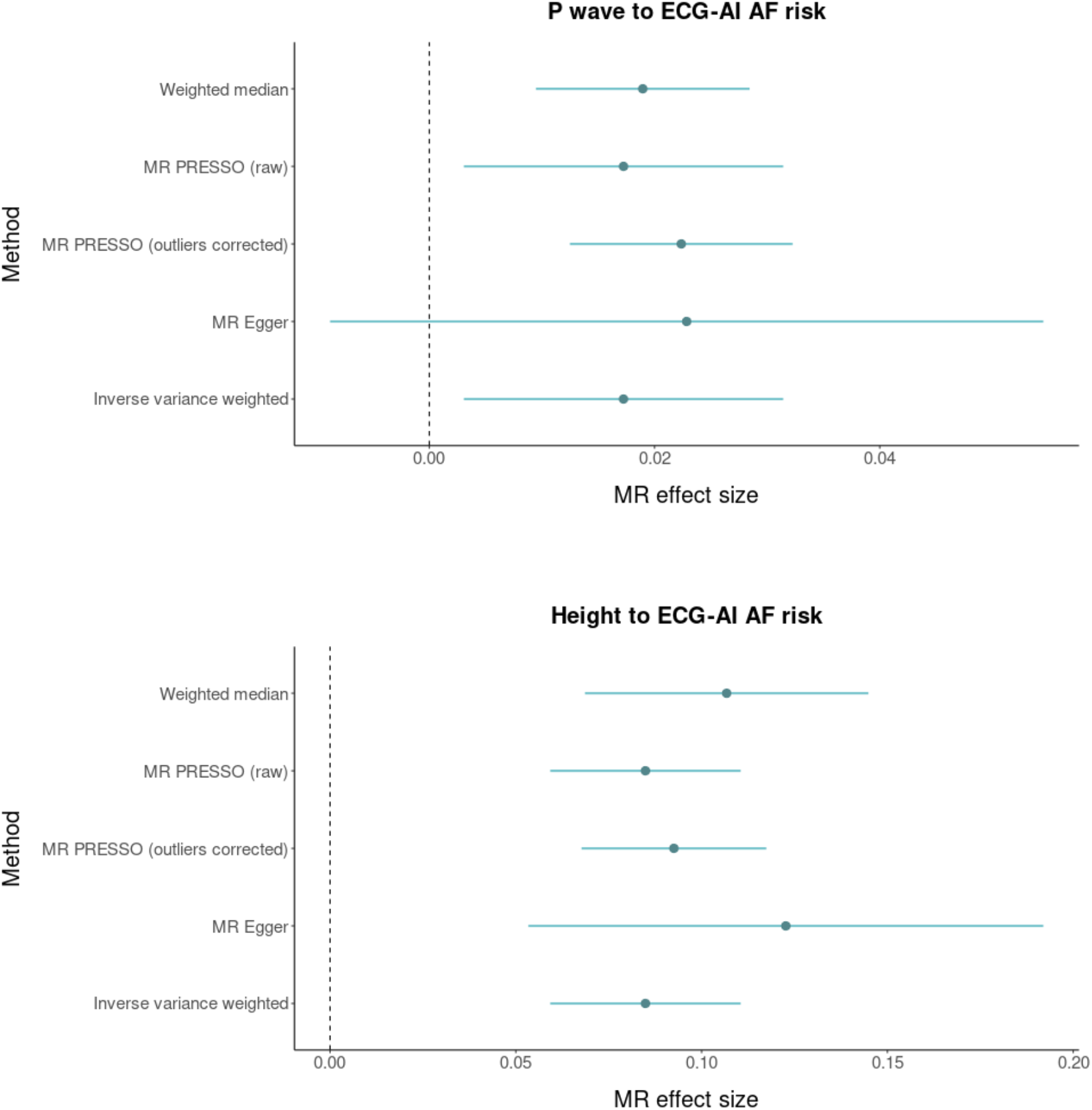
Mendelian Randomization analysis between P wave duration, body height, and atrial fibrillation (AF). The figure displays Mendelian randomization results assessing ECG P wave duration (top panel) and body heigh (bottom panel) for relations with genetically predicted ECG-AI risk. Mendelian randomization effect sizes are graphed on the x-axis. Dots represent point estimates and bars represent 95% confidence intervals. Dashed gray lines represent zero effect sizes. The five Mendelian randomization methods used in this analysis are shown in the y-axis.

## Discussion

We assessed the genetic basis of AF risk estimates generated from a validated deep-learning model using 12-lead ECGs. Despite the fact that individuals did not have AF at the time of ECG acquisition, we identified variants at three established AF susceptibility loci – *TTN, SCN5A*, and *SCN10A* – and at two novel loci that implicate body size measurements – *VGLL2* and *EXT1*. In contrast, our GWAS of CHARGE-AF derived AF risk did not identify any signals previously reported in a GWAS for AF, but identified loci linked to component clinical risk factors for AF that are included in the risk model. Our MR analyses provide supporting evidence that P wave duration and body height are causal risk factors for AF that mediate the ECG-AI risk estimates. Broadly, our findings imply that estimates of disease risk from deep learning models that use raw physiologic data, and risk scores more generally, are influenced by genetic susceptibility. In turn, such deep learning models may have the potential to identify individuals at risk for disease via specific genetic pathways.

Deep learning models of 12-lead ECGs for the identification of individuals with a high likelihood of AF have been reported^5–7^ but the interpretability and representations that underlie the risk estimates generated by the models have not been explained. We previously observed that ECG-AI estimates for AF risk are largely influenced by the P wave, a period corresponding to atrial depolarization and repolarization.^7^ Moreover, we have previously reported that both ECG-AI and clinical risk for AF are complementary.^7^ Here, we extend these observations by identifying genetic signals that have been associated with P wave duration,^36^ and documenting the distinct genetic profiles underlying risk estimates generated by ECG-AI and a clinical risk factor model.

Our findings have two major implications. First, risk estimates from the ECG-AI model are influenced by genetic mechanisms that are more specific to AF than are those from a clinical risk factor model. Specifically, loss-of-function variants in *TTN* have been associated with a substantially increased risk for AF^37,38^ and common variants at this locus have been associated with AF.^21^ *TTN* encodes titin, an integral protein involved in sarcomere development, structural integrity, and contractility.^39^ The ECG-AI GWAS also identified variation at the *SCN5A* and *SCN10A* loci. Both *SCN5A* and *SCN10A* encode the alpha subunits of voltage gated sodium channels and *SCN5A* is essential for myocyte depolarization. Genetic variants at these loci have been described in association with AF and ECG traits in prior GWAS^21,26,40,41^ and in rare familial forms of AF.^31,42–44^ *VGLL2* encodes for the vestigial like family member 2a protein, is critical for skeletal muscle development and contains a interacting domain for TEAD1, a member of the Hippo signaling pathway.^45^ *EXT1* encodes a protein involved in the production of heparan sulfate^36^, and has been implicated in the development of the outflow tract.^47^ We note that the *EXT1* and *VGLL2* loci are relative gene-dense loci, and we have focused here on the nearest genes as a means of prioritization.

In contrast, the genetic signals for CHARGE-AF risk were not specific to AF but reflected the diverse genetic mechanisms underlying the component risk factors in the model, including body height, body weight, blood pressure, and smoking status. Notably, the ECG-AI model loci linked to body size – *VGLL2* and *EXT1* – differ from the CHARGE-AF model loci linked to body size, implying that the manifestation of body size on the ECG may reflect different dimensions of body size from those measured conventionally using height and weight. The relative specificity of the ECG-AI model for genetic mechanisms underlying AF is further supported by our observation that the genetic correlation with AF from a prior GWAS was greater with ECG-AI predicted risk than with CHARGE-AF predicted risk.

Overall, our finding that predicted risk of AF from an ECG-based deep learning model is influenced by inherited susceptibility raises the possibility that an individual’s genetic predisposition to AF is inferrable using their raw ECG data alone, even prior to disease onset. Indeed, the GWAS of incident AF did not identify any significant genetic susceptibility loci, underscoring the power of performing a GWAS of models trained to predict risk of a disease, rather than of the disease itself. Future analysis is warranted to assess whether ECG-AI, and other risk models more broadly, can be used to specifically predict which individuals are predisposed to diseases via particular biological mechanisms. Such insights could theoretically have important implications for the personalization of prevention and therapeutics, identification of individuals with genetic disorders, and the use of deep learning or other risk models as digital biomarkers in addition to general risk prediction models.

Second, the genetic analysis of risk estimates generated by AI models may facilitate the interpretation of output, and underlying representations, of the models which may otherwise be difficult to interpret. Our GWAS, and MR analyses, highlight the fact that our ECG-AI model is influenced by heritable factors related to the P-wave and body size. The fact that ECG-AI can infer anthropometric traits^48^ suggests that deep learning model estimates from raw physiologic data can be influenced by numerous variables which manifest on the data modality under study. Height has been causally linked to AF risk previously.^34,35^ We propose the transfer of clinically-derived models to datasets with genomic information and subsequent genetic association testing, as a means to explore the factors which influence risk estimates from deep learning models.

We further note that whereas our approach focused on a single model task – the predicted risk of AF – examining the genetic architecture of the model latent space itself may reveal the genetic basis for the ECG representations learned by the model. As the number of samples with both ECG and genomic data increase, we anticipate greater statistical power to identify genetic signals underlying predicted disease risk, including rare large-effect variants. Moreover, we submit that the increasing availability of large-scale biobank data with raw data acquisition amenable to deep learning will enable examination of the genetic basis of other disease predictions. We anticipate that improved understanding of the relations between biological pathways and deep learning models may facilitate model application in clinical practice by enhancing clinician confidence in model outputs, and facilitating the inference of biological pathways that lead to disease risk in specific individuals.

Our study should be interpreted in the context of the design. First, our phenotyping algorithm used hospitalization and death records to ascertain disease status, which may lead to disease misclassification. However, before applying the ECG-AI model in UKBB, we omitted individuals that were not identified as prevalent AF by the phenotyping algorithm but were indicated as AF by their diagnostic statement accompanying ECG data, to reduce the impact of misclassification. Second, the MGH dataset we used to train the ECG-AI model and the UKBB dataset we used to perform genetic analyses were predominantly White British, which may limit the generalizability of our findings to populations of other ancestries. It is unclear if the results of the GWAS of ECG-AI reflect the underlying composition of the sample in which it was trained – future analysis of other ECG-AI models for AF prediction are warranted. Third, widespread replication of different model based GWAS will be necessary as additional and larger biorepository datasets emerge. Fourth, risk discrimination of the ECG-AI model in UKBB is moderate. A model with greater discrimination in the discovery sample may increase yield of genetic associations. Lastly, given roughly three years of follow-up after ECG in the UK Biobank, the power of our incident AF analysis may be limited.

In conclusion, we have shown that ECG-AI predicted AF risk reflects inherited predisposition to AF with a genetic background that is more specific for AF risk loci when compared to that for a clinical model. A polygenic risk score constructed using common variants associated with ECG-AI risk was significantly associated with incident AF in a prospective cohort. The interpretability of the ECG-AI model was improved by genetic analyses indicating that P wave duration and body height are likely to be two contributing factors forming the basis of AF risk predictions.

## Data Availability

All data produced in the present study are available upon reasonable request to the authors.

