## Supplemental Material for "Genetic Susceptibility to Atrial Fibrillation Identified via Deep Learning of 12-lead Electrocardiograms"

### **Supplementary Table 4. Summary statistics of the independent significant lead single nucleotide variants (SNVs) identified in the ECG-AI risk GWAS.**

| **SNVs in ECG-AI risk GWAS** | **Genome**  **Position (hg19)** | **Beta (se) in ECG-AI risk GWAS** | **P in ECG-AI risk GWAS** | **Beta (se) in CHARGE risk GWAS** | **P in CHARGE risk GWAS** |
| --- | --- | --- | --- | --- | --- |
| rs2627040 | 2-179601975 | 0.077 (0.014) | 4.55×10^−8^ | 0.004 (0.005) | 0.467 |
| rs41312411 | 3-38621237 | 0.055 (0.009) | 2.79×10^−10^ | -0.005 (0.003) | 0.172 |
| rs6801957 | 3-38767315 | -0.044 (0.006) | 2.68×10^−12^ | -0.007 (0.002) | 0.003 |
| rs9689288 | 6-117508467 | 0.043 (0.006) | 5.05×10^−12^ | 0.009 (0.002) | 2.97×10^−4^ |
| rs35186392 | 8-118857805 | 0.045 (0.008) | 7.22×10^−9^ | -0.0004 (0.003) | 0.897 |

### **Supplementary Table 5. Heritability and genetic correlation estimates for ECG-AI, CHARGE-AF, and previously reported AF GWAS.**

| **Traits** | ***h^2^* (GWAS sumstats)** | ***h^2^* (individual-level data)** |
| --- | --- | --- |
| ECG-AI risk | 7.0% (1.3%) | 13.0% (1.4%) |
| CHARGE risk | 24.3% (2.1%) | 36.5% (1.4%) |
|  | ***r_g_* (GWAS sumstats)** | ***r_g_* (individual-level data)** |
| AF and ECG-AI risk | 41.9% (7.3%) | 35.3% (13.7%) |
| AF and CHARGE risk | 28.2% (3.6%) | 18.9% (8.6%) |
| ECG-AI risk and CHARGE risk | 50.1% (7.4%) | 39.3% (4.5%) |

*h^2^*: heritability; sumstats: summary statistics; *r_g_*: genetic correlation. The previously reported AF GWAS summary statistics are derived from Roselli et al (2018).^1^

### **Supplementary Table 6. Polygenic risk and incident AF in a subset of White British participants (N=370,121).**

|  | **Individual models** | | **Interaction model** | |
| --- | --- | --- | --- | --- |
|  | **HR (95% CI)** | **P-value** | **HR [95% CI]** | **P-value** |
| PRS_ECG-AI_ | 1.08 (1.05-1.10) | 2.41×10^−9^ | 1.07 (1.04-1.10) | 1.29×10^−7^ |
| PRS_CHARGE-AF_ | 1.12 (1.09-1.15) | 3.57×10^−19^ | 1.11 (1.09-1.14) | 1.75×10^−17^ |
| Interaction term |  | | 1.00 (0.98-1.02) | 0.98 |

In the individual models, each PRS was tested separately as a predictor of AF, adjusting for age at enrollment, sex, genotyping array, and the first 20 principal components of ancestry. In the interaction model, main effect terms for each PRS were included along with a multiplicative interaction term between each PRS, adjusting for the above covariates. HR: hazard ratio; CI: confidence interval.

### **Supplementary Figure 1. LocusZoom plots of significant risk loci in the ECG-AI predicted 5-year AF risk GWAS.**

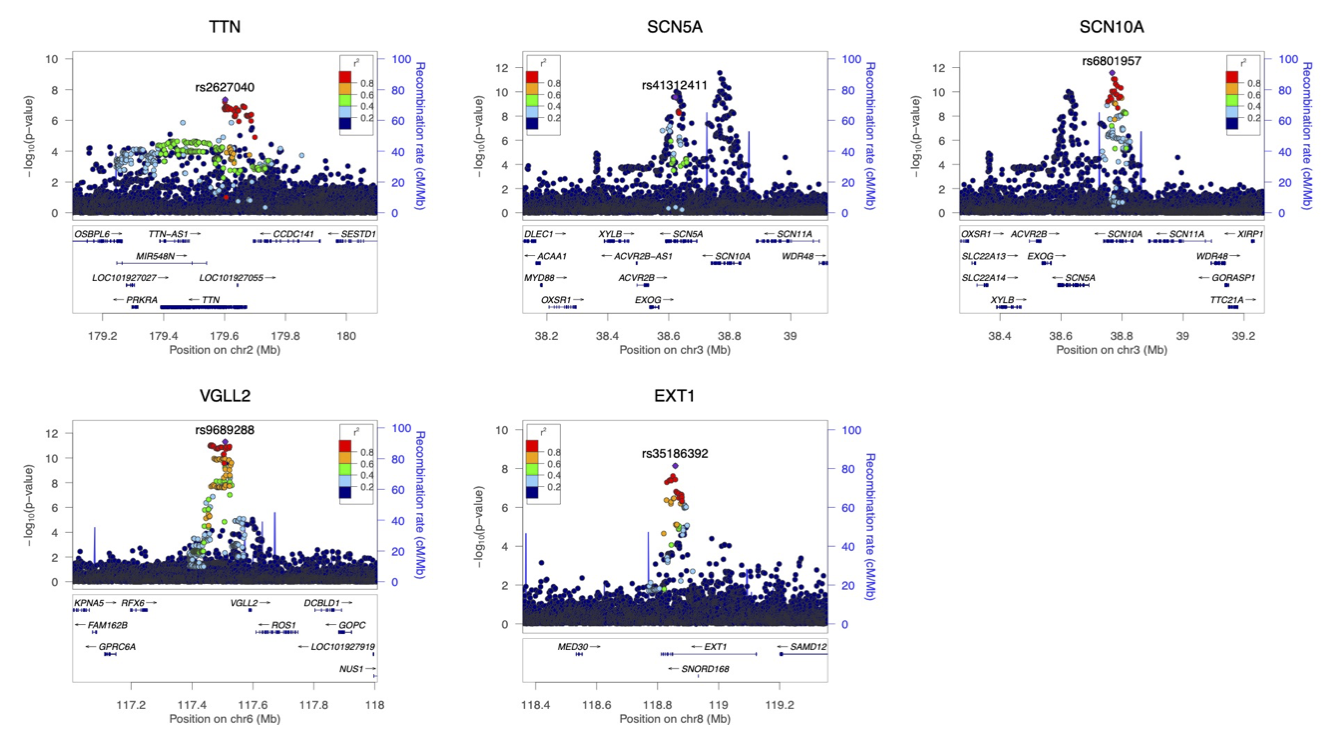

### **Supplementary Figure 2. LocusZoom plots of significant risk loci in the CHARGE-AF predicted 5-year AF risk GWAS.**

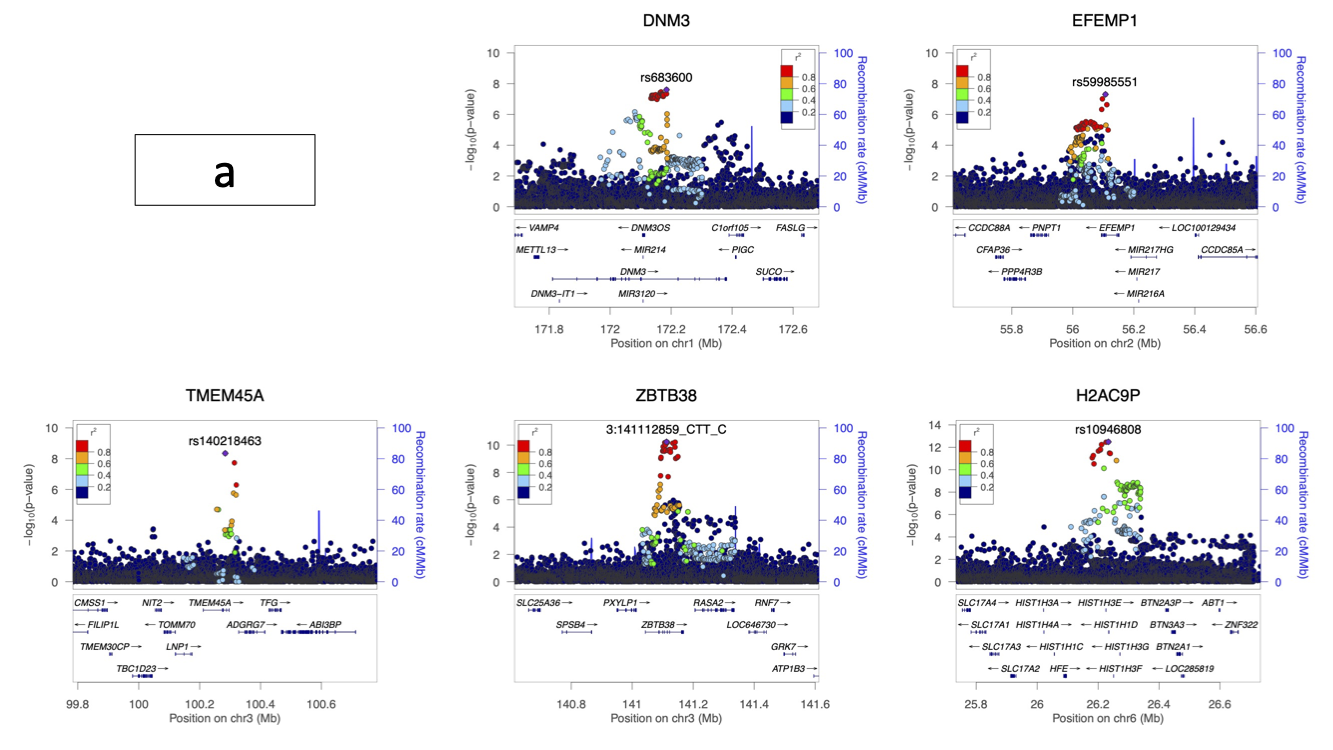

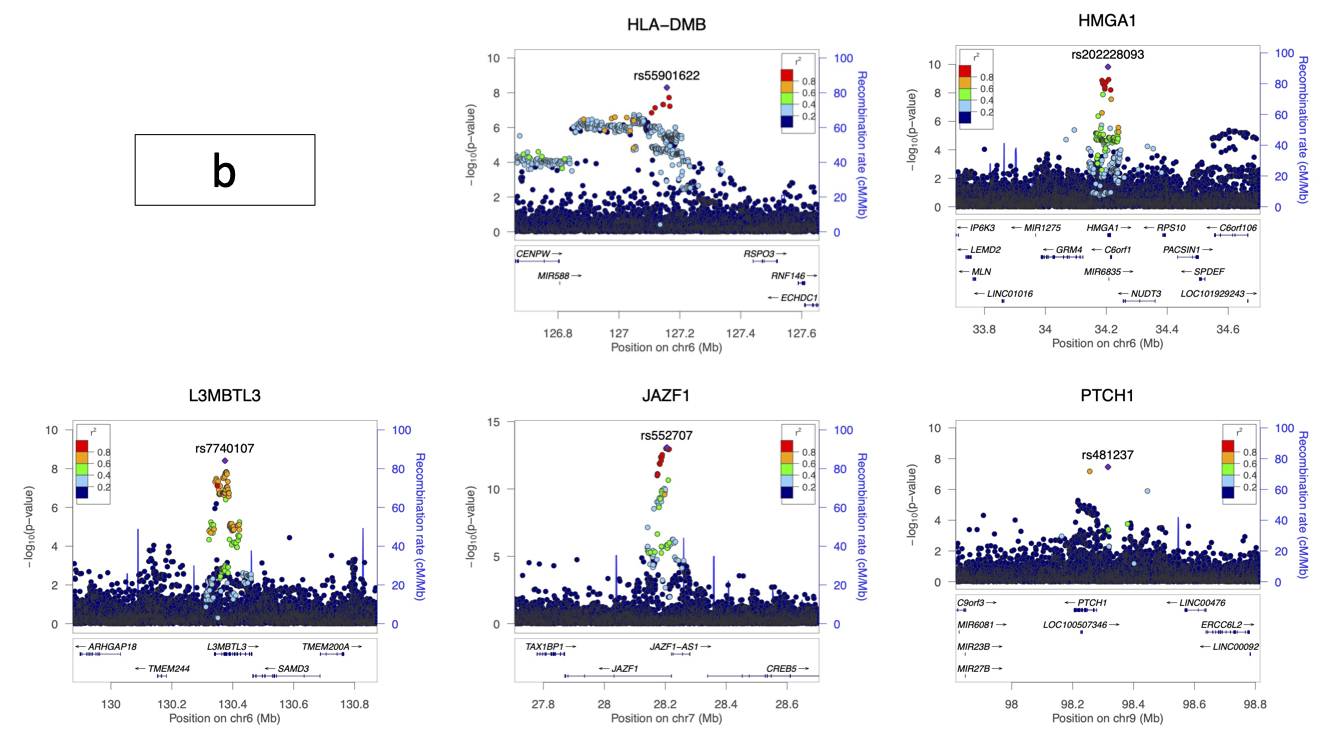

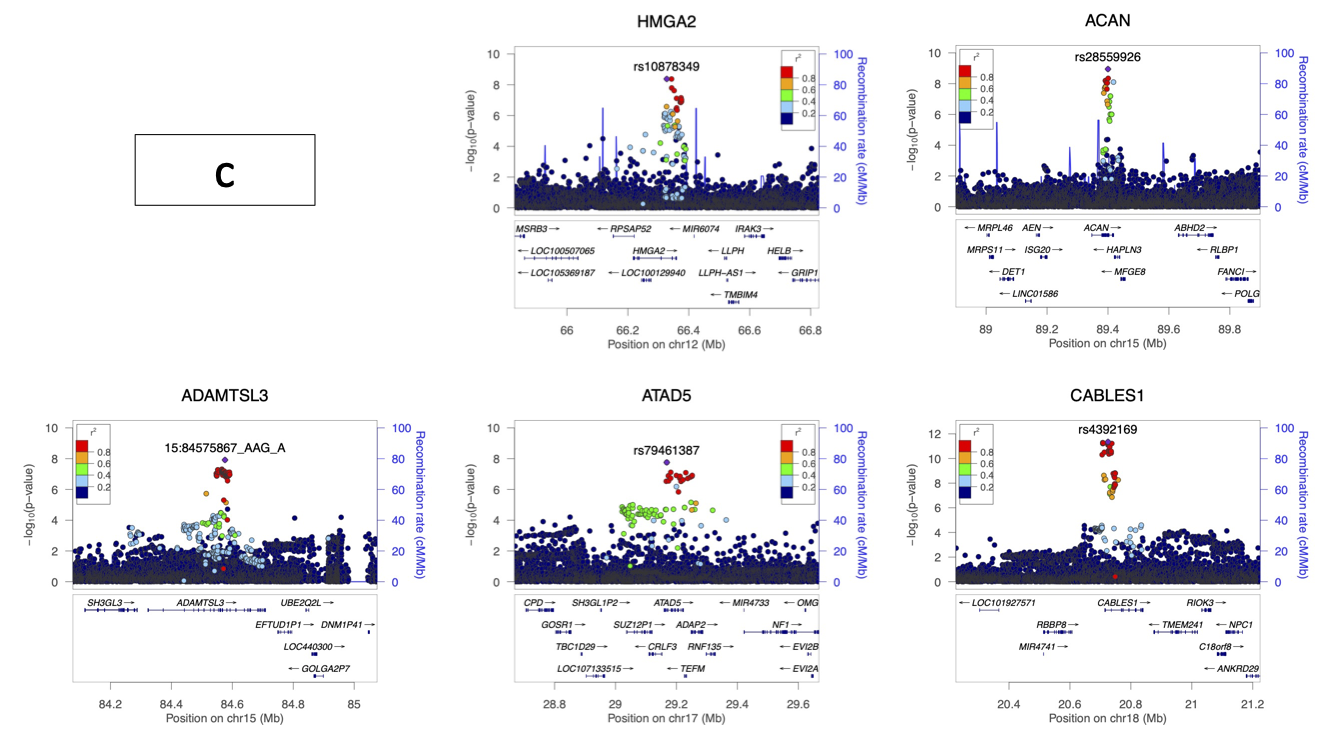

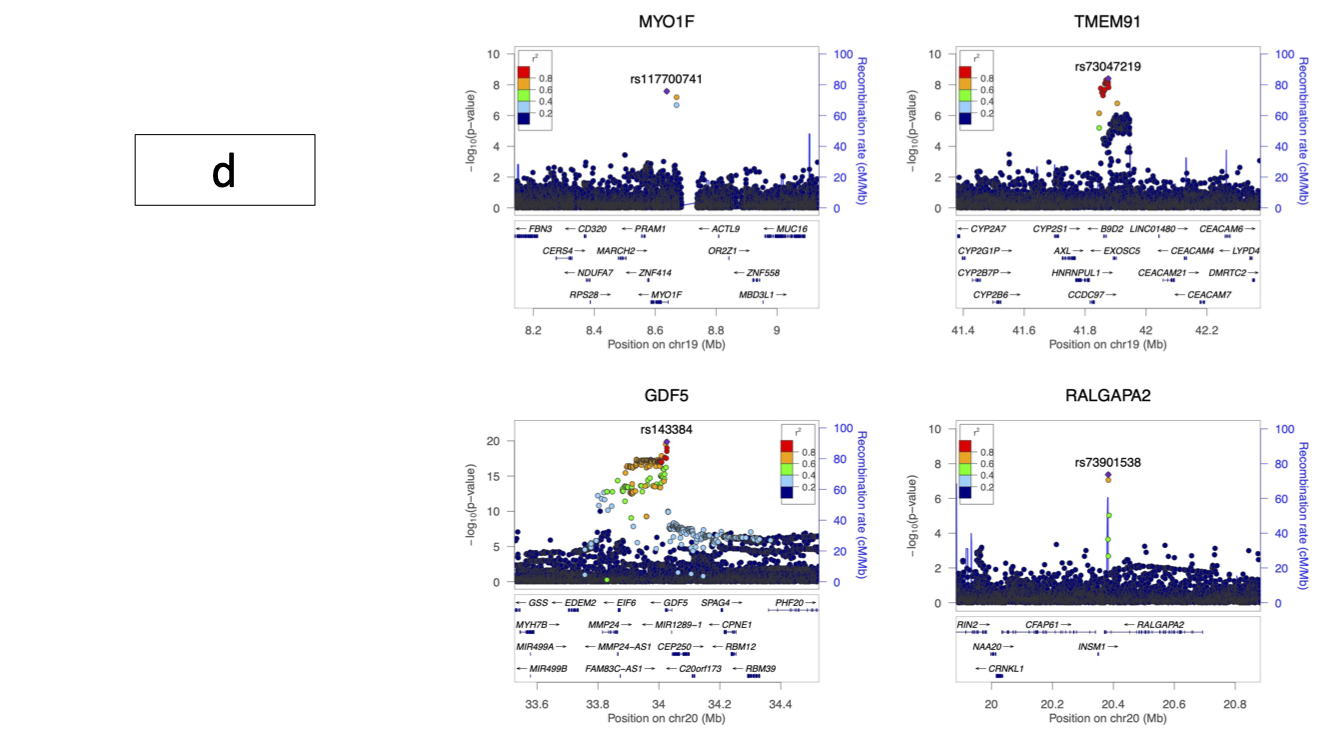

### **Supplementary Figure 3. ECG-AI and CHARGE-AF polygenic risk and cumulative incidence of AF.**

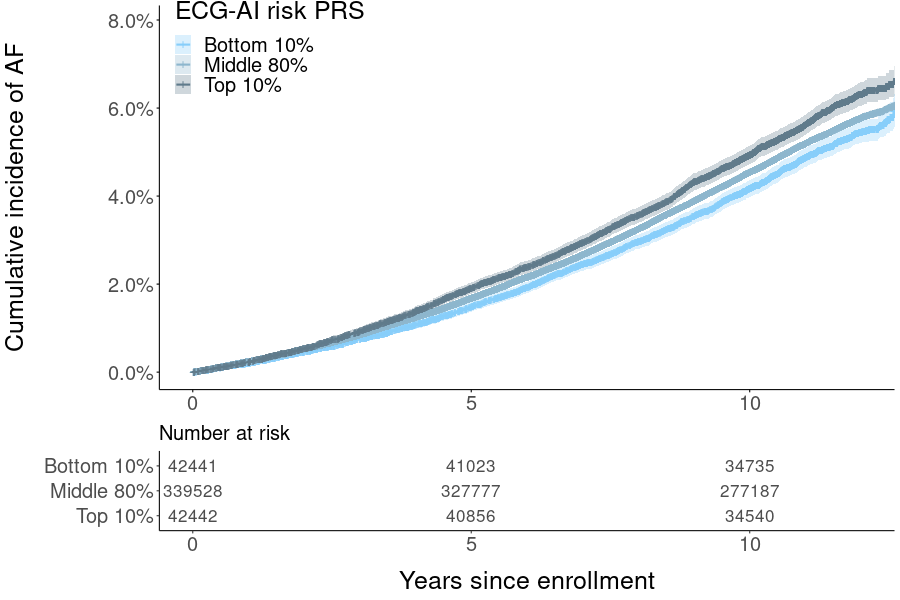

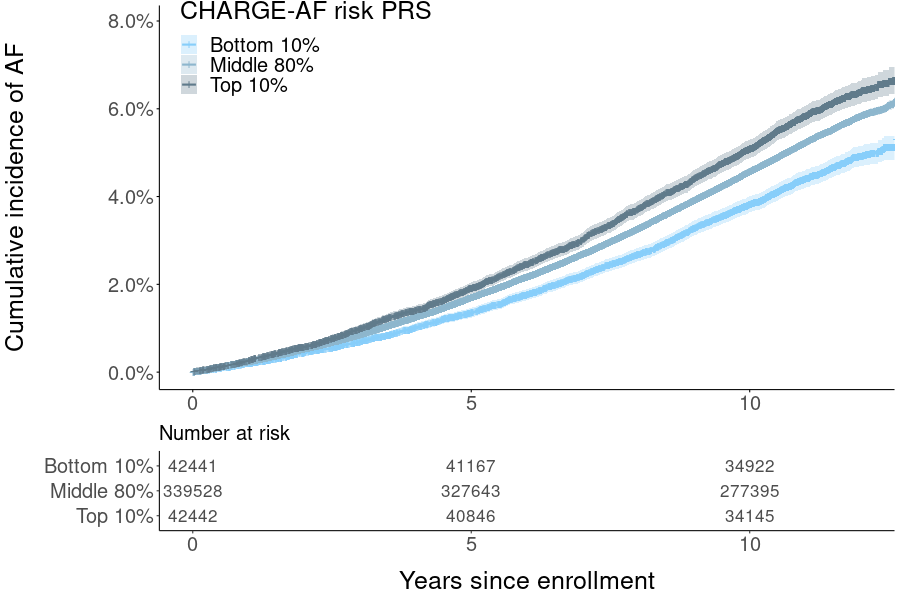

The cumulative incidence of AF is plotted for participants in three risk groups (lowest 10%, middle 80%, and highest 10% of the PRS distribution). Shaded regions around Kaplan-Meier survival estimates represent the 95% confidence intervals.

### **Supplementary Figure 4. ECG-AI and CHARGE-AF polygenic risk and cumulative incidence of AF in a subset of White British participants (N=370,121).**

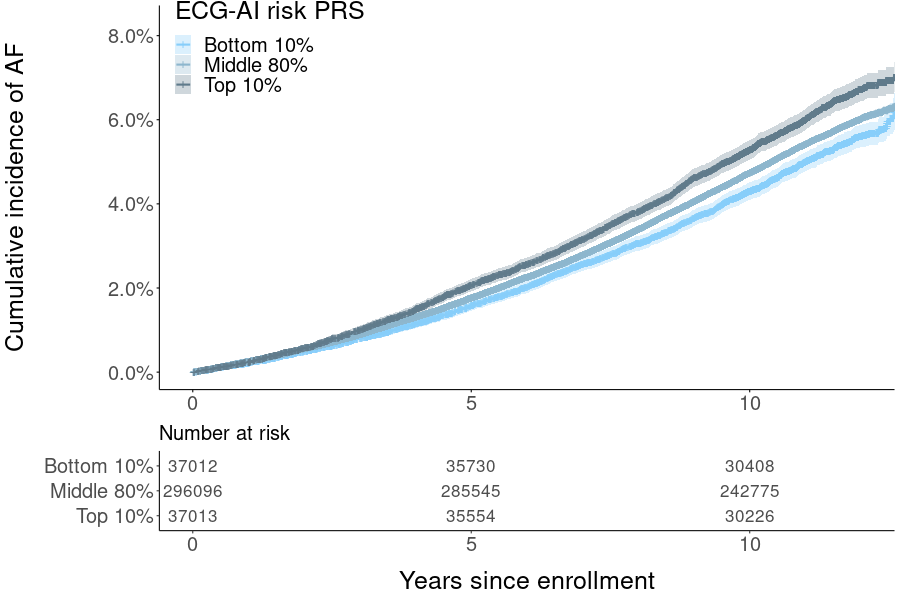

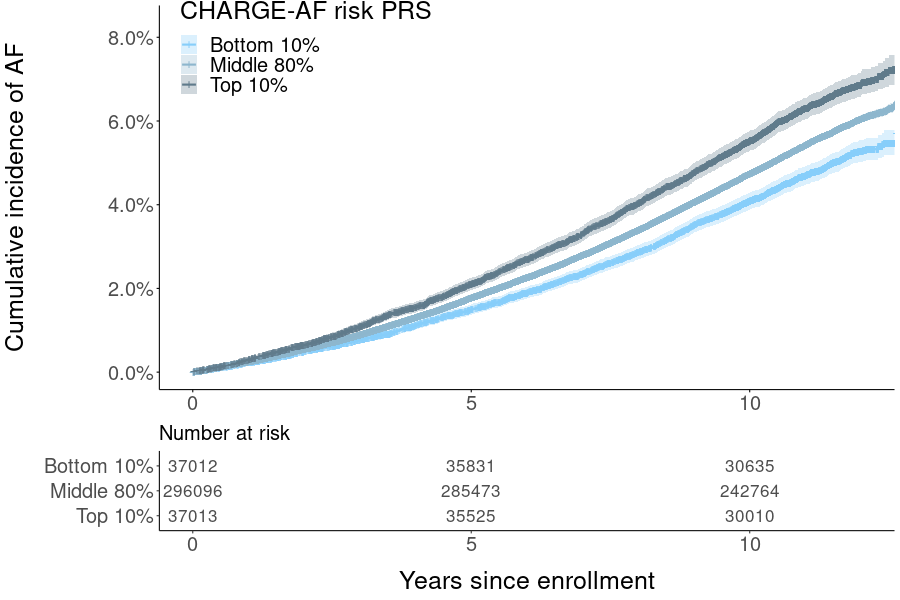

The cumulative incidence of AF is plotted for participants in three risk groups (lowest 10%, middle 80%, and highest 10% of the PRS distribution). Shaded regions around Kaplan-Meier survival estimates represent the 95% confidence intervals.
